# Adolescent-led sexual and reproductive health and rights research: qualitative evaluation of an innovative participatory action research project in Senegal

**DOI:** 10.1101/2021.08.26.21262685

**Authors:** Geneviève Fortin, Ashley Vandermorris, Mohamadou Sall, Britt McKinnon

## Abstract

Participatory approaches are increasingly popular in public health, but remain little used to address adolescent health issues. However, adolescent participation in research has enormous potential for identifying solutions to health issues that concern them. In Senegal, a youth-led participatory action research project was implemented to evaluate the potential of the approach to address adolescent sexual and reproductive health and rights issues. A qualitative evaluation was conducted in four Senegalese communities, where teams of adolescents were trained to conduct a research project and identify solutions relevant for their communities. Observations and interviews were carried out during results dissemination meetings in each of the communities. Based on participatory action research principles and expected adolescent participation outcomes, the evaluation of the project demonstrated the potential of the approach to identify relevant solutions, while promoting positive and meaningful adolescent participation. Despite some difficulties, such as community resistance, the youth researchers were able to successfully complete their research project, while developing their confidence and self-esteem. The adolescents were convinced that they could act as agents of positive social change. This project has shown that it is possible and relevant to involve adolescents in research projects, and that this approach has significant potential in global health.

## Introduction

Ensuring universal sexual and reproductive health and rights for adolescents is a global health priority (Hindin et al., 2016; United Nations, 2017). This includes high-quality sexual and reproductive health services and comprehensive sexuality education (CSE); the elimination of gender-based violence, child marriage and harmful practices like female genital mutilation/cutting (FGM/C); and the promotion of gender equality (Germain et al., 2015). Effective interventions to improve adolescent sexual and reproductive health and rights (ASRHR) are critical to achieve national health and development goals. Yet there remains a clear gap in the quality and availability of effective and scalable adolescent-focused interventions to improve ASRHR and growing recognition that the development of effective adolescent programming requires the involvement of adolescents themselves (Chandra-Mouli et al., 2015; Fatusi, 2016; World Health Organization, 2017). The involvement of adolescent girls in particular has been identified as an ethical and human rights imperative in the effort to achieve gender equity and universal ASRHR (World Health Organization, 2017).

Youth-led participatory action research (YPAR) is an innovative approach to research that engages adolescents in addressing issues that concern them (Ballonoff Suleiman et al., 2019; Lindquist-Grantz & Abraczinskas, 2020; Ozer, 2017; Ozer & Piatt, 2017; Villa-Torres & Svanemyr, 2015; World Health Organization, 2017). It is an approach to positive youth development that aims to promote equity and improve conditions that impact adolescents’ health, by respecting their fundamental right to participation (Ozer, 2017; Patton et al., 2016; UNICEF, 2018; Villa-Torres & Svanemyr, 2015; World Health Organization, 2017). Through direct participation in research, adolescents become agents of social and community change, while gaining opportunities for reflection and developing a deeper understanding of local issues (Ozer, 2017; Ozer & Piatt, 2017; Patton et al., 2016). In addition, this approach expands youth social networks and provides opportunities to develop skills in research, communication, collaboration and advocacy (UNICEF, 2018).

This mobilization of adolescents can help identify relevant solutions and interventions to change conditions that influence their health and well-being at the local level. It is from this perspective that we carried out an exploratory YPAR project in Senegal. This paper presents the evaluation of this project, which aims to determine the potential of YPAR to address ASRHR issues and identify implementation successes and challenges to inform future YPAR projects.

## Methods

### Qualitative approach

This evaluation was based on YPAR principles described by Villa-Torres and Svanemyr (2015) and on the Conceptual Framework for Measuring Outcomes of Adolescent Participation from UNICEF (2019) (Table I). The YPAR principles (column 1) guided the implementation of the project, and the evaluation therefore focused on how these principles were integrated. The adolescent participation outcomes (column 2) assess how well the YPAR process ensures meaningful participation of adolescents and respects their rights to participation. For this project, both types of outcomes were assessed using a qualitative descriptive approach and a content analysis, in order to reflect the perspective of the participants, remain close to the data, and inform subsequent practices (Caelli et al., 2003). The interview guides were built to assess each adolescent participation outcome and YPAR principle from three points of view: that of the adolescents involved as youth researchers, the youth researchers mentors, and key community members.

**Table I.** YPAR principles and adolescent participation outcomes used to guide the evaluation of the project, as described by Villa-Torres and Svanemyr (2015) and UNCEF (2019).

| **YPAR Principles** | **Adolescent Participation Outcomes** |
| --- | --- |
| Training and practice in research techniques | Sense of self-worth, self-esteem and self-efficacy |
| Practice strategic thinking and discuss strategies to influence change | Being taken seriously |
| Building support networks in collaboration with stakeholders | Making decisions |
| Power sharing between adults and young people in the process of research and action | Public and civil engagement |
| Power sharing among the young participants |  |
| Opportunities and advice for working in groups in order to achieve the expected objectives |  |
| Development of skills to communicate with other adults and young actors |  |

### Context

The YPAR project took place in the regions of Kaolack and Tambacounda, Senegal, between April and November 2019. These regions were selected to assess the feasibility of a YPAR approach among Senegalese populations from diverse socio-cultural backgrounds and in contexts with different ASRHR profiles. In terms of socio-demographic diversity and health status, the Kaolack region largely resembles that of the national population, while Tambacounda is one of the most disadvantaged regions in Senegal (see Table II).

**Table II.**
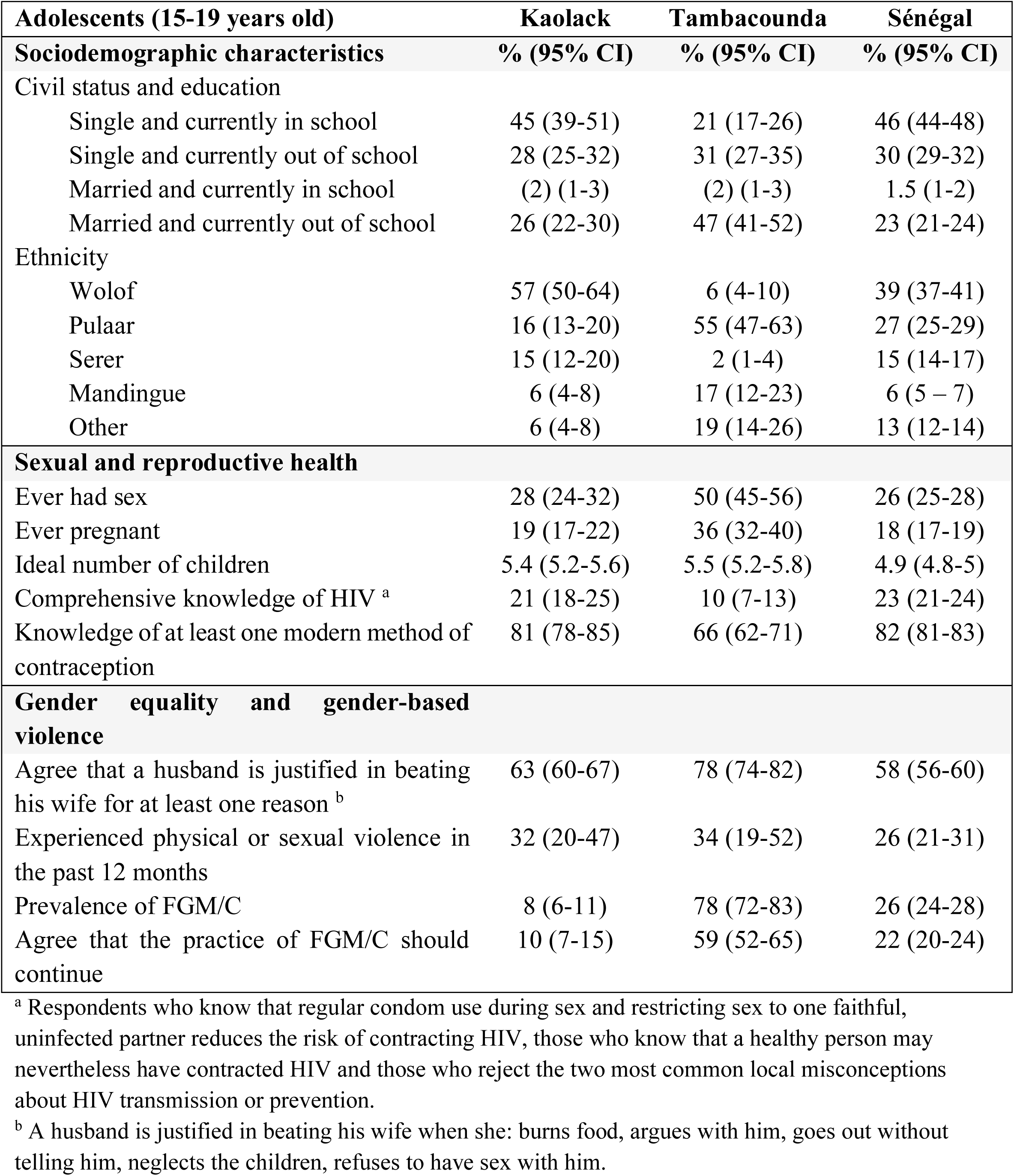
Adolescent sexual and reproductive health and rights indicators for the regions of Kaolack and Tambacounda, Senegal (authors’ calculations using Senegal 2017 Demographic and Health Survey data).

### YPAR design

From these regions, 4 communities (2 rural, 2 urban) were selected to host youth-led research teams supported by an adult mentor, following discussions between the study team, NGO partners and local government and health authorities. To ensure confidentiality for participants, the communities of Kaolack will be identified by A1 and A2, and those of Tambacounda as B1 and B2, A1 and B1 being rural and A2 and B2 urban. Each team included 3 to 5 youth researchers 18-21 years old, and one mentor. Adolescents were selected based on age, gender, school level and other factors to ensure diversity within each team. The communities were involved in the selection process, and adolescents were identified by local key community members. Potential adolescent participants were interviewed by the study team to confirm their interest and motivation to join the project. All mentors were female and had master’s level training in a social science research discipline. All teams followed a participatory research process as delineated in Figure 1:

**Figure 1.**
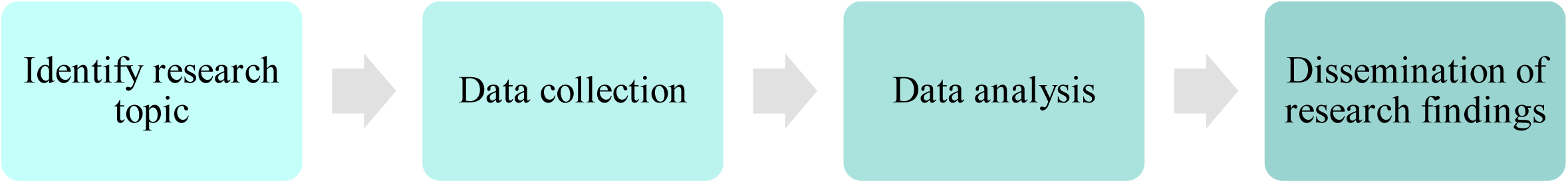
Participatory action research process as followed by the adolescents involved on this project.

The initial training for the YPAR project was conducted by a team of Senegalese and Canadian study investigators from April 23 to 25, 2019. It took place in two stages: the mentor training followed by the youth researcher training. The mentor training focused on PAR methods, mentor responsibilities in the project and how to ensure proper follow up throughout the study. The training for youth researchers introduced the principles of YPAR, including methodology, data collection and analysis, and their responsibilities as researchers. Both trainings included an overview of ASRHR issues in Senegal and emphasized the importance of research ethics.

Supervision visits were conducted by the Senegalese study co-investigator throughout the project to evaluate the progress made by teams in preparing their research, in collecting and analyzing data, and writing reports. In addition, a mid-term workshop took place in August 2019 to assess the progress of all teams, and for them to share their work with their colleagues and receive feedback from the research team.

### YPAR process

During their training, youth were introduced to the fundamentals of research methodology, a number of data collection methods, including quantitative (questionnaires, surveys) and qualitative (semi-structured interview, focus group discussion (FGD), PhotoVoice), and ethics. As a first research step, each team determined priority ASRHR issues in their community by reviewing available literature and conducting informal interviews in the community. They used this information to select their research question and decide on a methodological approach for the research project (Table III). Ultimately, 2 groups decided to proceed with a qualitative project using semi-structured interviews and FGDs, while 2 developed mixed methods studies using questionnaires followed by FGDs or semi-structured interviews.

**Table III.** Titles of the projects led by adolescents by community.

| Community | Title |
| --- | --- |
| A1 (rural) | Determinants and psychosocial consequences of teenage pregnancies |
| A2 (urban) | Determinants of early marriage among adolescents in A2 |
| B1 (rural) | Study on early marriage in the Tambacounda region: case of the rural commune of B1 |
| B2 (urban) | The frequency of STIs in adolescents aged 15 to 21 in the B2 district |

After conducting their research, all youth researcher teams presented their findings and recommendations at community dissemination meetings in November 2019. The project evaluation took place during those dissemination meetings, where it was possible to learn more about their research project; conduct interviews with youth researchers, mentors and community members; and assess the potential of the YPAR projects to tackle ASRHR issues.

### Evaluation of YPAR

#### Data collection

Semi-structured interviews were conducted with youth researchers (YR), mentors (M) and community members (CM) between November 23 and 26 2019, following dissemination meetings in the 4 communities. Only youth researchers with a sufficient level of French were selected for the interviews (n=6 of 16 YR), and all mentors were interviewed (n=4). Community members who were involved in or exposed to the project, and who attended the dissemination meeting in their community, were selected according to their availability for an interview following the meeting (n=14; 11 men, 3 women). All interviews were audio recorded with the consent of the participants. The interviews with the youth researchers and mentors were carried out and transcribed in French by a Canadian qualitative researcher, while interviews with community members were conducted by Senegalese master’s students in local languages, then translated and transcribed in French. All interviewers were previously unknown to the participants involved on the project. Direct observations were also conducted during the dissemination meetings by members of the study team.

#### Data analysis

All transcribed documents were coded using Atlas.ti qualitative analysis software v.8.4.0 (922) (Scientific Software Development GmbH, Berlin, Germany). Content analysis was performed based on the adolescent participation outcomes and YPAR principles. Initial coding was conducted using descriptive, emotional and structural codes, followed by a focused coding phase to group the codes according to the different evaluation criteria. The field observation notes were used to enrich and validate the results obtained from the interview analysis.

#### Rigour and reflexivity

Memos were written to document the analytical process, and peer debriefing allowed for validation of the results by all collaborators (Caelli et al., 2003). All authors believe adolescents have a right to participation and that creating spaces for them in their communities is a key factor to reduce health inequalities. We acknowledge that ASRHR is a sensitive and complex topic that can be influenced by cultural and community factors. Researchers have different cultural references from the participants and come from various backgrounds. These postures influence the interpretation of the data.

#### Ethical considerations

This study was reviewed and approved by the Research Ethics Board at the Hospital for Sick Children in Canada, and the National Committee for Health Research in Senegal.

## Results and findings

### PAR principles

#### Training and practice in research techniques

The youth researchers identified that the training effectively enabled them to acquire new knowledge and develop research skills, with a particular emphasis on confidentiality: “*I learned how to talk to people. And to get consent before registering them and writing their names and keeping it a secret from others. I learned a lot about confidentiality*.*”* (YR; A1)

Overall, the training seemed to have been a source of motivation for most adolescents, although some limitations were raised, for example that the training was too short (5 days) and condensed. They would have preferred to have more time to better assimilate the information. Others mentioned the lack of background information received on their research topic, such as papers or reports, which presented a barrier when analyzing their data, as stated by this mentor: ‘*Sometimes, to be able to argue, to be able to support the words of our interviewees, we needed documents for better… to have readings on our subject. So, we didn’t have these documents, especially with regard to our research topic. We really needed documentation, yes, especially on our subject. Because we are working on the frequency of STIs*.*’* (M; B2)

#### Practice strategic thinking and discuss strategies to influence change

In each community, adolescents developed recommendations based on their findings, which were presented to their community during dissemination meetings. This community outreach strategy to raise awareness about the issue was positively endorsed during the interviews with community members, as well as youth researchers. However, even though they recognized the value of the project, some community members questioned its real impact. Indeed, it seems like there was concern that the proposed solutions by adolescents would be ignored, as stated by this community member who was wondering about next steps: ‘*I got a lot of information during their presentation. I didn’t have the option of having them. So, having them now will also allow me to have new axes of intervention, or new acts that will allow me as a member of the community to take them into account in certain interventions that I am used to do with my peers. Talk to them about it. What actions must we take to be able to get out of this?*’ (CM; A1)

From the adolescents’ perspective, most believed they could have real impact in their community to bring change, as stated by this participant when asked what advice they could give to other adolescents: *‘[…] I can tell him*, “You, as a youth researcher, can participate in the development of society, or you can participate in eradicating this disease.” *You understand? For example, I can tell her, “*If you work on teenage pregnancies, when you educate people, those who will be aware of what you are saying, it can decrease teenage pregnancies.*” So, I can tell him that he can participate, him. That is to say that he, as a young person, he has a very important place, he must not see himself as “*I am young, I am a teenager, I am… I am not considered; I cannot serve society.” *He has to get to the idea first: I can change that! For example, “*There is this problem, I am young, I can change it.” *You understand?’* (YR; B2) Youth researchers have thus recognized that they can be agents of change, through awareness-raising, discussion and community participation.

#### Building support networks in collaboration with stakeholders

Youth researchers and mentors were able to develop their support network through various meetings with key actors from their community, with the project partners and among themselves. Several mentioned that they were able to get help and gain knowledge from the people met through the project, including mayors, imams, doctors, nurses, midwives, and village chiefs. This adolescent discussed this newfound support network, and how their community has helped them gain insight on health issues: ‘*For example, health providers, traditional healers, and adolescents too, the main targets, they too have given us support. It’s because compared to them, we learned other things that we didn’t know. You understand? That is to say that we, when we went into the field, it was from their knowledge that we really got to know this problem. This is the problem that adolescents face in this locality*.’ (YR; B2)

The adolescents also raised the issue of providing support for their community, which reinforces their sense of empowerment: ‘In the neighborhood where we had to do the study, people, when they see us, if they have any questions in this area, can ask us. So before, if we passed each other, it was just a greeting and then that’s it. But now, if they have any problems, they can come to us and ask, thinking that we can point them in the right direction since we have had training in this regard. […] It is true that we have a certain notion about the matter, but the doctors are more qualified. So even if you want to answer some of these questions, you can only answer some, but others you can tell them to go see the doctor and things.’ (YR; B2)

#### Power sharing between adults and young people in the process of research and action

Youth researchers were confronted with potential participants refusing to collaborate during data collection. In these situations, the power dynamic between adolescents and adults was difficult for them. In communities with traditional and conservative values, the hierarchy of power is very important and taboo subjects like ASRHR can be difficult to tackle, especially for youth.

‘*[Adolescents don’t] have that much power, no. Because usually, and even to make decisions, they have to call us, say, “Can I do this?” Sometimes you encourage them to go on, but they’re afraid to take initiatives or do things. Well, I understand, because this is the first time they’ve done this sort of thing, so it’s understandable*.’ (M; A2)

‘*Others refused to cooperate, others told us it was taboo, others said… that is, they see it as a teenage influence. That is to say, here, the religious especially consider it a taboo subject, it is when we talk about sex*.’ (YR; B2)

‘*Sometimes adults and kids alike refuse to talk to us. Because they are not aware of the point. […] And many adults, like young people, refused the discussion. But most of them agreed to speak with us, showed us the way, advised us to continue*.’ (YR; A2)

However, adolescents mentioned that they felt respected by the adult researchers with whom they worked and were able to share their knowledge with them. Being able to interact with adults on a foundation of equality and respect helped build their confidence: *‘[Our relationship] was fair. Because they listened to us, they paid attention to what we said, if there were any corrections, they corrected us. They weren’t belittling us by saying “We are superior to them, they have no knowledge of the matter, so we are not going to listen to them, or we are not going to take into account what they are going to say*.*” Instead, they were able to support us, put us in the right conditions to make us feel comfortable*.’ (YR; B2)

#### Power sharing among the youth participants

The power sharing dynamics between the youth researchers was characterized by equality, mutual aid and respect. The 4 youth-led research teams included girls and boys, and all mentioned the equal relationship between genders and the advantages of working in mixed groups, as stated by this adolescent: ‘*Yeah, mixed group is good. Because you may not have the same ideas, so girls can have better ideas than boys, or boys can have better ideas than girls. So, we will put them together to be able to find the best [ideas]*.’ (YR; B1)

Mentors worked to ensure sufficient participation from the various youth researchers in their group. This seemed more difficult in groups where some youth had lower education level. On the other hand, the mentors and the youth researchers with more education mentioned having provided support to their less educated colleagues to ensure their opinions and ideas were also taken into account. All participants agreed that youth with less education should take part in the project, as their participation could be particularly beneficial, as stated by this adolescent who was not currently enrolled in school: ‘*We were all equal. […] This project made a difference*.*’* (YR; B1)

#### Opportunities and advice for working in groups in order to achieve the expected objectives

In terms of working as a group, collaboration and commitment were mostly used to describe the youth researchers’ experiences.

‘*To work well in a group, […] I think it’s the same as other jobs, you have to be serious, and say that it’s a job that will give me an opportunity in the future. You have to be serious and, above all, you have to be committed. Because if we’re not engaged, we don’t want to do well, we don’t want to do anything. You have to be committed, say that this is an opportunity for the future. Being young, you have to contribute to the good improvement of your community*.’ (YR; A2)

It seemed that for some, daily chores, family responsibilities and school represented barriers to their consistent involvement in the project.However, their youth researcher colleague were understanding towards such family and school responsibilities.

‘*There are times when I am not available. Because in my house, I am… It is I, alone, who manage the field work, and in this case, we must meet on evenings to do work on the project, and for that I cannot come and do the work with youth researchers. You have to not come, to go to the fields. […] It is in this case, like me, I make absences in the project*.’ (YR; A1)

#### Development of skills to communicate with other adult and young actors

Interviewed adolescents and mentors all said that their communication skills improved over the course of the project, whether for public speaking, approaching strangers, expressing themselves clearly and presenting their ideas, or improving their French. However, language barriers were an important challenge for many, especially for less educated adolescents who did not understand French or were not fluent in Wolof, the most common language in Senegal, although it is not the dominant language in all regions. A mentor explained how it could be a challenge, and how they overcame it: ‘*During the training there were no translators, mainly in the Mandingue language. [They had difficulty] keeping up, because we spoke Wolof during the youth training, and they did not understand Wolof well. Because there are some of them who don’t understand French too. Because there are young people with less education. […] It was a little complicated. Even during the data collection too, the translation of some French words was also a problem. We overcame these problems because there were strategies in place [*…*] We had someone, he is here today, he understands French better, he also understands Mandingue. He could translate some words that were complicated. We could ask him at any time to translate words that are a little difficult for youth*.’ (M; B1)

The above quote shows an interesting example of how adolescents got support from their community. Overall, communication skills have been developed and the experience has been positive, as stated by this adolescent, who presented their results in front of her community: ‘*Because even before, in class, to participate I was shaking. Even during training, in Kaolack, to introduce myself, say my name, I was shaking. The others were telling me*, “No, gotta talk”, *they encouraged me and things, to come here like that. But over time, I adapted. There is an improvement in the meantime. I was proud today*.’ (YR; B2)

From a community perspective, the adolescents involved in the project are now seen as being able to communicate with other young people on important issues: ‘*There are some advantages, because, first of all, it was the young people who got involved and it allowed them to know and to make others known. These youth researchers, what they learned, they pass on to their comrades at school, at home and in the neighborhoods*.’ (CM; B2)

### Adolescents Participation Outcomes

#### Sense of self-worth, self-esteem and self-efficacy

The youth researchers expressed great pride in having participated in the project, especially to have been able to help their community. Developing new skills helped increase their self-confidence and make new connections: *‘[My participation] made a difference, because before I took part in this project, I didn’t have that much knowledge to be able to identify these problems. But during this project, I learned a lot and I know a lot of people now too*.’ (YR; B1)

Some adolescents reflected that they now see themselves as researchers and expressed a desire to pursue future research projects. During the dissemination meetings, a few youth researchers who had left school expressed a desire to resume their studies. This was also a key point for this mentor: ‘*The benefits are mostly with young people. So at least they learned to do research. Some have told me, “*Ma’am, we wanted to go back to class just because of this research.” *Because that’s interesting, these are benefits. Really, youth who have dropped out of school, but who, because of this project, want to return to school. It makes me very happy. So they felt a bit of the interest in studies and research. Others, they say to you*, “Ma’am, me, later, I will do research.” *To show that they really learned something, they liked it, and it’s nice. And also to see, there are efforts that have been made, young people who did not speak, but in the project, they became more comfortable talking with people, breaking into their community. And it’s nice*.’ (M; A1)

Additionally, some youth researchers expressed an enhanced sense of self-efficacy. They felt that they could now bring solutions to the issues they care about, and that even though they are young, they have the capacity to help their communities.

‘*I got knowledge that I didn’t have, because I didn’t know there were so many problems in the community. And the project let me know that all these issues are in my community. […] If someone asks me one day “*What are the problems in your community?*”, it will be really easy to list them and also provide solutions to these problems*.’ (YR; A2)

The feeling of self-efficacy described by adolescents also seems to be challenging some of the traditional perceptions of the role of youth within communities. Indeed, some adolescents expressed newfound capacity to help their community: ‘*Through this study, I saw that even if I am young, I can be of service to my community. I’m young, but that doesn’t mean that I always have to be there, sit back – I can be useful to my community, help and when I have knowledge, share my knowledge too*.’ (YR; B2)

#### Being taken seriously

During dissemination meetings, it was evident that community members took the work of youth researchers seriously. The issues chosen by adolescents aligned with community concerns and local ASRHR priorities. In addition, the community’s openness to listen to youth researchers during dissemination meetings reinforced their feelings of being taken seriously.

‘*I think the community will make changes, since a lot of people encourage us and congratulate us for participating in this training, in this project. I think the population will be aware tomorrow, I think so*.’ (YR; A2)

However, during interviews with community members, some respondents seemed to have a different perspective, questioning the relevance of adolescents in research: ‘*If they were adult researchers, it could have changed a lot of things, like the community health workers who are there. There are some people who do not want to discuss with the little ones. […] [If the adults] leave their concerns at home to come and raise awareness, all people will follow them, while for the young people, some will say to themselves* “These young people, they only waste our time.”’ (CM; B1)

This participant presents his own positive perception of the involvement of young people in the projects, but nonetheless suggests that some members of the community who attended the dissemination meeting questioned the validity of the data presented by the adolescents: ‘*An adult researcher will not be able to chat with a young person. But if it is a young person doing the study, there will indeed be a conversation between young people because the subject is about young people. […] At the data level, after the presentation, there are people who say that some data is unreliable, just as the PowerPoint presentation is not well done*.’ (CM; A1)

#### Making decisions

According to the adolescents involved in the project, decision-making came mainly from the mentors or adults involved in the project. The youth researchers shared their opinions and exchanged ideas, but also expressed discomfort with making decisions. The participants identified collaboration and mentoring as more important than autonomy in decision making.

‘*We respected each other, we agreed on everything we will do. Not one person will decide, if our mentor tells us to do this, we all do it. It’s not somebody who’s going to say* “No, I won’t do it”, *or two people. No, we all do it together*.’ (YR; B1)

Even though decisions were mostly made by mentors, adolescents were the ones who chose the issue they worked on, and they conducted their project themselves. From their mentors’ perspective, they had power and their voices mattered: ‘*Adolescents*… *They have the capacity and are equipped too, through their training, so that they can reflect on the sexual and reproductive health problems suffered by adolescents in the locality as well. […] They are the ones who carried out the research project. They were the ones who conducted the research project, they had power, yes, over adults*.’ (M; B1)

#### Public and civic engagement

The commitment of youth researchers to their communities was evident: all expressed pride that they were able to contribute to the improvement of the chosen issues. Raising awareness appeared to be an important factor motivating their engagement with their communities: ‘*Yes, things will change. With the support of others, youth researchers, we could change our lives. People the same age as us, yes… After educating parents about the issues, to keep young girls in school and tell young girls to know their values and prepare for their future*.’ (YR; A1)

‘I can feel proud and say that I have brought something to [my community]. I have been useful to my community in this regard. So, the more I can help them, the better.’ (YR; B2)

While the issues identified by the adolescents were recognized as important by all participants, including community members, participants also reflected that relations between adolescents and adults remain unequal and taboos are very difficult to overcome. A participant reinforced this point by explaining that young people still cannot speak with their parents, but that the project has nevertheless helped reduce taboos: ‘*The taboo has diminished today compared to yesterday. Now we are talking about it, even if it is not between young people and their parents, but between young people it is done. [*…*] To tell you that the taboo subject is no longer relevant, even if it leaves some traces*.’ (CM; A1)

Despite difficulties associated with taboos, youth researchers expressed a commitment to bringing changes to their communities beyond the study: ‘*Sometimes you want to talk with people, and it will be difficult to make them aware, sometimes they even don’t want to talk with you. And that is, really, is a disaster. Because people who don’t want to speak for their own sake don’t know what to expect. It’s really hard working with these people. But I made a decision to continue, even if the project stopped, to do awareness days*.’ (YR; A2)

## Discussion

This evaluation brought to light many positive elements of YPAR, confirming the potential of this methodology and supporting its use to address ASRHR issues. Our assessment of YPAR principles and adolescent participation outcomes revealed that adolescents can significantly benefit from being included in research, even if some challenges arise. A key success was the engagement of girls in research teams, where they were able to participate in mixed groups, without being oppressed. All the girls played key roles within their team, including presenting the results in their community. This suggests significant potential for YPAR to create safe and inclusive environments for young girls, a critical element to consider in ASRHR programs (Santhya & Jejeebhoy, 2015). While adolescents faced some resistance from their communities, this is not surprising. It is well established that social norms can be a barrier to the success of YPAR projects (LPC Consulting, 2012), reflecting the historically recognized resistance of societies to mobilizing adolescents (UNICEF, 2018).

The testimonies of the youth researchers involved in this YPAR project in relation to each of the YPAR principles were largely positive. Despite some challenges encountered, adolescents expressed that participating in YPAR was an engaging and enriching experience. In all communities, youth were faced with resistance and persistent taboos surrounding sexuality. In Senegal, these taboos represent a major barrier to health education activities, both at the family and community level (Jourdan et al., 2010). These taboos may also explain the limited access to information about sexually transmitted infections. Despite this, the youth researchers showed great openness and motivation to improve the sexual and reproductive health of their communities. Unanimously, they were all ready to get involved again in such projects and were all convinced of their capacity to promote change and put in place solutions to ASRHR issues. Indeed, the participation of adolescents in research, as agents of social and community change, can transform traditional models of care and influence the implementation of interventions from various cultural perspectives, in addition to allowing the study of sensitive health topics, including ASRHR (LPC Consulting, 2012; Ozer, 2017; Ozer & Piatt, 2017; Patton et al., 2016).

The youth researchers in this study developed a great sense of confidence and self-esteem, some even describing themselves as a researcher at the end of the project. They expressed pride in the work they accomplished and felt able to help their community. YPAR can indeed foster adolescent participation in innovative ways, involving them at every step of the research process and overcoming many barriers that persist in other forms of ASRHR participatory projects (Fakoya et al., 2021). ASRHR programs often involve limited engagement of local leaders (Santhya & Jejeebhoy, 2015), however, through this project, adolescents felt empowered to work in collaboration with important community actors to make decisions about ASRHR, creating a more supportive environment. More so, the youth researchers’ engagement in their communities was evident: their desire to continue working beyond this project to improve and raise awareness was a testament to this commitment.

The dissemination meetings were essential to ensure meaningful youth participation, as it is a key element of a YPAR project (Freshwater, 2005), providing a space for them to be heard on important topics. During the meetings, adolescents had the opportunity to speak in front of elders, including their parents and members of their communities, which is not a common practice in Senegal. Their participation to this YPAR project gave them legitimacy to do so, enabling them to be heard by their community. Even though resistance emerged from some community members, the audience listened to the youth and showed engagement and interest in the project. *Space, Voice* and *Audience* are three of the essential features identified by UNICEF to ensure meaningful participation (UNICEF, 2018). Youth researchers’ desire to raise awareness contributes to deconstructing taboos and paves the way to find relevant solutions. Adolescents are thus real agents of social change in their community, contributing to their collective well-being.

The social ecology in which a YPAR takes place, including social and community norms, influences the level of participation of adolescents (UNICEF, 2018; World Health Organization, 2017). Unequal power dynamics and community hierarchies are barriers to the success of YPAR initiatives and can particularly disadvantage young girls (Iwasaki et al., 2014; Patton et al., 2016; Santhya & Jejeebhoy, 2015; UNICEF, 2018). Socio-cultural contexts may help explain certain elements of community and even participant resistance. In Senegal, young people are often raised to leave decision-making to the elders. Despite this, adolescents in this study indicated that they believe they can bring change within their communities, that adolescents matter and have a crucial role in addressing ASRHR issues. Thus, this project supports the potential and relevance of using a YPAR approach, promoting adolescents inclusion even in contexts where their participation challenges social norms.

Overall, this evaluation demonstrated an active participation in the project, an involvement valued by communities, and the development of individual and community empowerment. This is consistent with previous findings that adolescents’ empowerment and their influence on their communities can arise from meaningful participation in PAR (UNICEF, 2018). In this project, adolescents were actively involved from the start, thus enriching individual and community empowerment capacities. Their desire to pursue ongoing community outreach strategies reflects their sense of their power to influence change. From a critical global health perspective, this perception of greater power is particularly relevant and suggests a transformation of unequal power dynamics and greater equity in the research process, a central aspect of YPAR (Ozer et al., 2020).

This evaluation has limitations, including a possible courtesy bias that may have arisen despite all participants being encouraged to share their experiences and comments, positive or negative, with honesty. Furthermore, one of the main limitations of the evaluation is that it did not take into account the experience of youth researchers who did not complete the project. Likewise, youth researchers with an insufficient level of French could not be interviewed, and their experience was not taken into account. The growing interest in YPAR in public health is reflected through global actions, including guidelines from the World Health Organization’s Global Accelerated Action for the Health of Adolescents (AA-HA!) (World Health Organization, 2017). This framework encourages adolescent participation in research that concerns them, promoting an approach to work together, ‘WITH adolescents, FOR adolescents’. In addition, AA-HA! recommends continuing local consultations to promote the acceptability of projects aimed at improving adolescent health, in particular by consulting parents or legal guardians. Conducting local consultations is a step that could have been implemented for this project and may have contributed to a greater acceptability. Local consultations could improve adults’ understanding of the project and its acceptability, and leave more room for adolescents to make their own decisions, even though it is non-traditional for their communities.

## Conclusion

This YPAR project was exploratory and aimed to identify key elements to consider for future projects focusing on ASRHR. This evaluation demonstrates the great potential to tackle ASRHR issues in a low- and middle-income settings, as well as the benefits for adolescents who have no previous experience in research. This project shows that it is possible to include adolescents in global health research projects and to support them in examining solutions in their own community, despite local challenges. The positive and meaningful overall experience endorsed by youth participants confirms the relevance of further exploring YPAR projects as an innovative solution to ASRHR issues.

## Data Availability

The data collected through this research is not available to protect participants' identity, confidentiality and safety.

## Acknowledgments

The authors would like to thank Adama Touré, Abdoulrazak Bagourmé, Aminata Dior Ndiaye, and Rudy Broers of Plan International’s Strengthening Health Outcomes for Women and Children (SHOW) programme in Senegal and Dr Stanley Zlotkin, Dr. Diego Bassani and Katie McLaughlin (SickKids Centre for Global Child Health) for their support and technical inputs. We acknowledge the regional and district medical offices in Kaolack and Tambacounda for their collaboration and support during the development and execution of the study. Finally, we extend our appreciation to the mentors, youth researchers, and community participants for their dedication, enthusiasm and participation in this study.

